# Seroprevalence of Antibodies to SARS-CoV-2 among Health Care Workers in Kenya

**DOI:** 10.1101/2021.03.12.21253493

**Authors:** Anthony O. Etyang, Ruth Lucinde, Henry Karanja, Catherine Kalu, Daisy Mugo, James Nyagwange, John Gitonga, James Tuju, Perpetual Wanjiku, Angela Karani, Shadrack Mutua, Hosea Maroko, Eddy Nzomo, Eric Maitha, Evanson Kamuri, Thuranira Kaugiria, Justus Weru, Lucy B. Ochola, Nelson Kilimo, Sande Charo, Namdala Emukule, Wycliffe Moracha, David Mukabi, Rosemary Okuku, Monicah Ogutu, Barrack Angujo, Mark Otiende, Christian Bottomley, Edward Otieno, Leonard Ndwiga, Amek Nyaguara, Shirine Voller, Charles Agoti, David James Nokes, Lynette Isabella Ochola-Oyier, Rashid Aman, Patrick Amoth, Mercy Mwangangi, Kadondi Kasera, Wangari Ng’ang’a, Ifedayo Adetifa, E. Wangeci Kagucia, Katherine Gallagher, Sophie Uyoga, Benjamin Tsofa, Edwine Barasa, Philip Bejon, J. Anthony G. Scott, Ambrose Agweyu, George Warimwe

**Affiliations:** KEMRI-Wellcome Trust Research Programme, Kilifi, Kenya; KEMRI Center for Infectious and Parasitic Diseases Control Research, Alupe, Kenya; Kilifi County Hospital, Kilifi, Kenya; Department of Health, Kilifi County, Kenya; Kenyatta National Hospital, Nairobi, Kenya; Alupe Sub-County Hospital, Busia, Kenya; Institute of Primary Research, Nairobi, Kenya; Kocholia Sub-County Hospital, Busia, Kenya; Busia County Referral Hospital, Busia, Kenya; Department of Health, Busia County, Busia, Kenya; Department of Infectious Diseases Epidemiology, London School of Hygiene and Tropical Medicine, UK; Ministry of Health, Government of Kenya, Nairobi, Kenya; Presidential Policy and Strategy Unit, The Presidency, Government of Kenya; Nuffield Department of Medicine, Oxford University, UK

**Author notes:** Corresponding author: Anthony O. Etyang, 813, Mbuyuni Bldg, P.O. Box 230-80108, Kilifi, Kenya. Equal contribution (Ruth Lucinde, Henry Karanja, J. Anthony G. Scott, Ambrose Agweyu, George Warimwe).

**Keywords:** SARS-CoV-2, Health Care Workers, Antibodies, Seroprevalence

## Abstract

**Background:** Few studies have assessed the seroprevalence of antibodies against SARS-CoV-2 among Health Care Workers (HCWs) in Africa. We report findings from a survey among HCWs in three counties in Kenya.

**Methods:** We recruited 684 HCWs from Kilifi (rural), Busia (rural) and Nairobi (urban) counties. The serosurvey was conducted between 30^th^ July 2020 and 4^th^ December 2020. We tested for IgG antibodies to SARS-CoV-2 spike protein using ELISA. Assay sensitivity and specificity were 93% (95% CI 88-96%) and 99% (95% CI 98-99.5%), respectively. We adjusted prevalence estimates using Bayesian modeling to account for assay performance.

**Results:** Crude overall seroprevalence was 19.7% (135/684). After adjustment for assay performance seroprevalence was 20.8% (95% CI 17.5-24.4%). Seroprevalence varied significantly (p<0.001) by site: 43.8% (CI 35.8-52.2%) in Nairobi, 12.6% (CI 8.8-17.1%) in Busia and 11.5% (CI 7.2-17.6%) in Kilifi. In a multivariable model controlling for age, sex and site, professional cadre was not associated with differences in seroprevalence.

**Conclusion:** These initial data demonstrate a high seroprevalence of antibodies to SARS-CoV-2 among HCWs in Kenya. There was significant variation in seroprevalence by region, but not by cadre.

## INTRODUCTION

Health care workers (HCWs) are critical in the acute-care response to epidemic waves of COVID-19, but they are also required to sustain normal health services beyond COVID-19. HCWs are considered to be at high risk of infection with SARS-CoV-2^1^. It is unclear whether the seroprevalence of SARS-CoV-2 antibodies among HCWs is more closely associated with community or hospital-based transmission risk as indicated by professional cadre. In some hospitals, seroprevalence was higher among cadres in lower paid jobs with little patient contact (e.g. housekeepers, porters) suggesting the source of infection may be their crowded living conditions rather than occupational risk^2^. Because the overwhelming majority (>90%) of infections in Kenya are asymptomatic^3^, and because PCR testing of nasal and oropharyngeal (NP/OP) swabs is challenging, the true extent of infection in this group has been difficult to determine in Kenya^4^ and, indeed, in most low and middle-income countries (LMICs).

Serological surveys can estimate cumulative incidence of SARS-CoV-2 infection in either key groups, such as HCWs, or the general population^5^. They can also assess the effectiveness of infection prevention and control measures, which is important in sub-Saharan Africa (sSA) where the availability of personal protective equipment and other preventive measures is constrained. To date HCW serosurveys in sSA have been limited to urban hospitals^6-8^; there are no surveys from rural hospitals, where resources are even more constrained. Serosurveys on different population groups or in different geographical regions can also inform vaccine prioritization policies. This is especially important in LMICs where only a small proportion of the population are likely to receive vaccines in the early phase of the vaccine campaign^9^.

Because the presence of antibodies to SARS-CoV-2 appears to be strongly protective against repeat infection over a 6-month period^10, 11^, knowledge of past infection could be useful for avoiding unnecessary quarantines which would help preserve the limited numbers of personnel available to deal with the pandemic and other health needs in the region.

We report initial findings from SARS-CoV-2 antibody testing from HCWs in three sites in Coastal, Central, and Western Kenya.

## METHODS

### Study sites and Participants

Study sites (Figure S1) were selected after consultation with the individual county COVID-19 Rapid Response Teams (RRTs). For Kilifi County, a predominantly rural area located on the Indian Ocean coast, we enrolled participants at Kilifi County Hospital, which is the main referral facility in the region. For Busia County, which is also predominantly rural and located in the western region of Kenya, we enrolled HCWs at Busia County Referral Hospital, the main referral facility in the area, and two other facilities in the county; Alupe Sub-County Hospital which has been designated as the isolation facility for COVID-19 patients in the county, and Kocholia Sub-County Hospital. In Nairobi County, the capital city of Kenya, we enrolled HCWs at the Kenyatta National Hospital (KNH), the main referral facility for the city as well as the country^12^.

We used a variety of strategies to recruit a convenience sample of HCWs at each of the study sites, including word of mouth, advertising at hospital notice boards and messages sent via mobile phone. HCWs of all cadres were eligible to participate in the study. In Kilifi and Busia we aimed to recruit ≥ 50% (N=441) of the 882 HCWs working in the healthcare facilities, which we considered to be both feasible and likely to provide a representative sample. We used a slightly different strategy at KNH, where the primary aim of the study was to determine incidence and antibody kinetics among HCWs. With a target sample size of 180 at KNH, it would not have been possible to get a representative sample because of the difficulty in obtaining updated lists of the ∼5,000 HCWs present at the hospital who are either directly employed by the hospital, the University of Nairobi and/or the Kenya Medical Training College, or are trainees from these and other institutions from Kenya and overseas^12^. Here, we aimed to recruit HCWs from different departments of the hospital, who were likely (by self-report) to be available for a year-long longitudinal study of incidence and antibody kinetics.

### Ethics and Consents

Serosurveillance was conducted as a public health activity requested by the Kenya Ministry of Health and ethical approval for publication of these data was obtained from the Kenya Medical Research Institute Scientific and Ethics Review Unit (KEMRI/SERU/CGMR-C/203/4085). HCWs provided written and/or verbal informed consent for participation in the study. Results of the antibody testing were reported confidentially to each HCW together with information explaining the implications of the test results.

### Sample collection and processing

The study took place between 30^th^ July 2020 and 4^th^ December 2020. Data collection was performed by members of staff from the participating hospitals, trained on the study procedures.

We collected 6ml of venous blood in sodium heparin tubes from each participant. Serum was obtained by centrifuging the samples at 450 x g for 5 minutes before storage at –80°C. Samples were then transported in dry ice to the KEMRI-Wellcome Trust research laboratories in Kilifi for assays.

A simple one-page questionnaire (provided in the appendix) was administered to the HCWs either electronically or on paper, in which data on demographic and clinical characteristics were collected.

### ELISA for SARS-CoV-2 Spike Protein

All samples were tested at the KWTRP laboratories in Kilifi for IgG to SARS-CoV-2 whole spike protein using an adaptation of the Krammer Enzyme Linked Immunosorbent Assay (ELISA)^13^. Validation of the assay was described previously^14^. Briefly, sensitivity, estimated in 174 SARS-CoV-2 PCR positive Kenyan adults in Nairobi and a panel of sera from the UK National Institute of Biological Standards and Control (NIBSC), was 92.7% (95% CI 87.9-96.1%); specificity, estimated in 910 serum samples from Kenya drawn in 2018 (i.e. pre-pandemic period), was 99.0% (95% CI 98.1-99.5). Results were expressed as the ratio of test OD to the OD of the plate negative control; samples with OD ratios greater than two were considered positive for SARS-CoV-2 IgG.

### Statistical methods

We assumed that the seroprevalence at each of the study sites would be 5-10%. We estimated that a minimum of 180 participants per site would generate prevalence and/or seroconversion (at KNH) estimates with a precision of ± 0.05 to ±0.10 for the site-specific estimates and a precision of ±0.01 to ±0.03 for the overall prevalence.

Continuous variables were summarized as means and standard deviations if normally distributed and medians with interquartile ranges for non-normally distributed variables. Categorical data were presented as counts and percentages. Bayesian modelling was used to adjust seroprevalence estimates for the sensitivity and specificity of the assay. Non-informative priors were used for all parameters, and the models were fitted using the Rstan software package^15^ (see appendix for code). We tested for associations between seroprevalence and professional cadre and site, respectively using multivariable logistic regression.

All analyses were conducted using Stata™ Version 15 software (College Station, Texas, USA) and R version 3.6.1 (Vienna, Austria).

## RESULTS

We recruited 684 HCWs from Nairobi, Busia and Kilifi (Figure 1 and Table 1). The numbers of the HCWs that we recruited as a proportion of total number of staff at the facilities were 70% in Kilifi, 50% in Busia, and ∼4% in Nairobi. The mean age ± SD of the participants was 35 ± 11 years and 54% were female. Sixteen (2%) of the HCWs reported that they had acute respiratory symptoms at the time of sample collection.

**Table 1:** Characteristics of study participants.

| Site, Dates of sample collection | All Sites<br>30 Jul-04Dec<br>N=684 |  | Kilifi<br>13 Oct-04 Dec<br>N= 200 |  | Nairobi<br>30 Jul-25 Aug<br>N=183 |  | Busia<br>19 Oct-23 Oct<br>N=301 |  |
| --- | --- | --- | --- | --- | --- | --- | --- | --- |
| Characteristic | n | (%) | n | (%) | n | (%) | n | (%) |
| Female | 372 | 54 | 113 | 57 | 99 | 54 | 160 | 53 |
| Age group (years)<sup>a</sup> |  |  |  |  |  |  |  |  |
| 18-30 | 232 | 34 | 65 | 33 | 67 | 37 | 100 | 33 |
| 31-40 | 226 | 33 | 69 | 35 | 54 | 30 | 103 | 34 |
| 41-50 | 117 | 17 | 34 | 17 | 31 | 17 | 52 | 17 |
| 51-60 | 85 | 13 | 20 | 10 | 20 | 11 | 45 | 15 |
| >60 | 17 | 3 | 8 | 4 | 9 | 5 | 0 | 0 |
| PCR swab previously collected | 250 | 37 | 31 | 16 | 77 | 43 | 142 | 47 |
| Positive on previous swab | 5 | 2 | 1 | 3 | 0 | 0 | 4 | 3 |
| Symptoms at sample collection | 16 | 2 | 0 | 0 | 16 | 9 | 0 | 0 |
| Chronic illness<sup>b</sup> | 18 | 3 | 0 | 0 | 18 | 10 | 0 | 0 |
| Work in Covid Unit<sup>c</sup> | 50 | 7 | 0 | 0 | 0 | 0 | 50 | 16 |
| Cadre |  |  |  |  |  |  |  |  |
| Nurse | 152 | 22 | 42 | 21 | 50 | 27 | 60 | 20 |
| Doctor | 85 | 12 | 21 | 11 | 53 | 29 | 11 | 4 |
| Clinical Officer | 79 | 12 | 48 | 24 | 4 | 2 | 27 | 9 |
| Support staff<sup>d</sup> | 117 | 17 | 33 | 17 | 21 | 11 | 75 | 25 |
| Pharmacy | 19 | 3 | 4 | 2 | 7 | 4 | 8 | 3 |
| Laboratory | 64 | 9 | 13 | 7 | 7 | 4 | 44 | 15 |
| <sup>e</sup>Other | 168 | 25 | 39 | 20 | 41 | 22 | 76 | 29 |

<sup>a</sup>Age missing for 7 individuals<sup>b</sup>Chronic illness- hypertension, diabetes, asthma, HIV<sup>c</sup>None of the HCWs in Nairobi and Kilifi worked in a Covid unit<sup>d</sup>Support staff includes: kitchen staff, patient porters, security staff, records clerks<sup>e</sup>Other staff includes-hospital administrators, supervisors, cashiers, accountants

**Figure 1:**
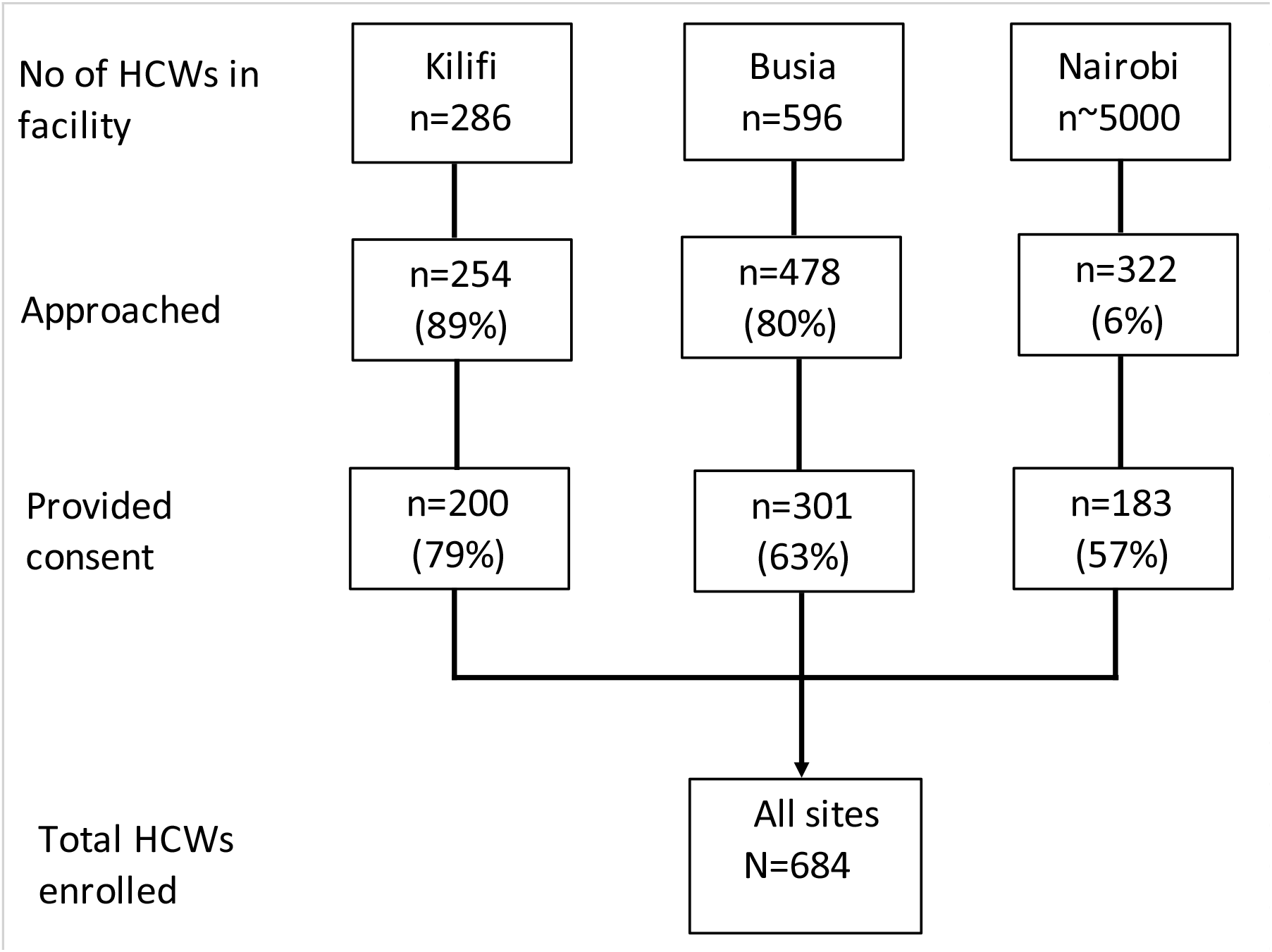
Study flow chart.

Out of the 684 HCWs, 135 (19.7%) were seropositive for antibodies to SARS-CoV-2 (Table 2). After adjusting for test performance characteristics, the seroprevalence was 20.8% (95% CI 17.5-24.4). Adjusted seroprevalence among the different cadres ranged from 12.5% (95% CI 5.4-21.8) among Clinical Officers to 34.2% (95% CI 23.7-45.8) among doctors. There was a higher seroprevalence among HCWs in Nairobi (43.8%, 95% CI 35.8-52.2) compared with Kilifi (11.9%, 95% CI 7.2-17.6) and Busia (12.6%, 95% CI 8.8-17.1).

**Table 2:**
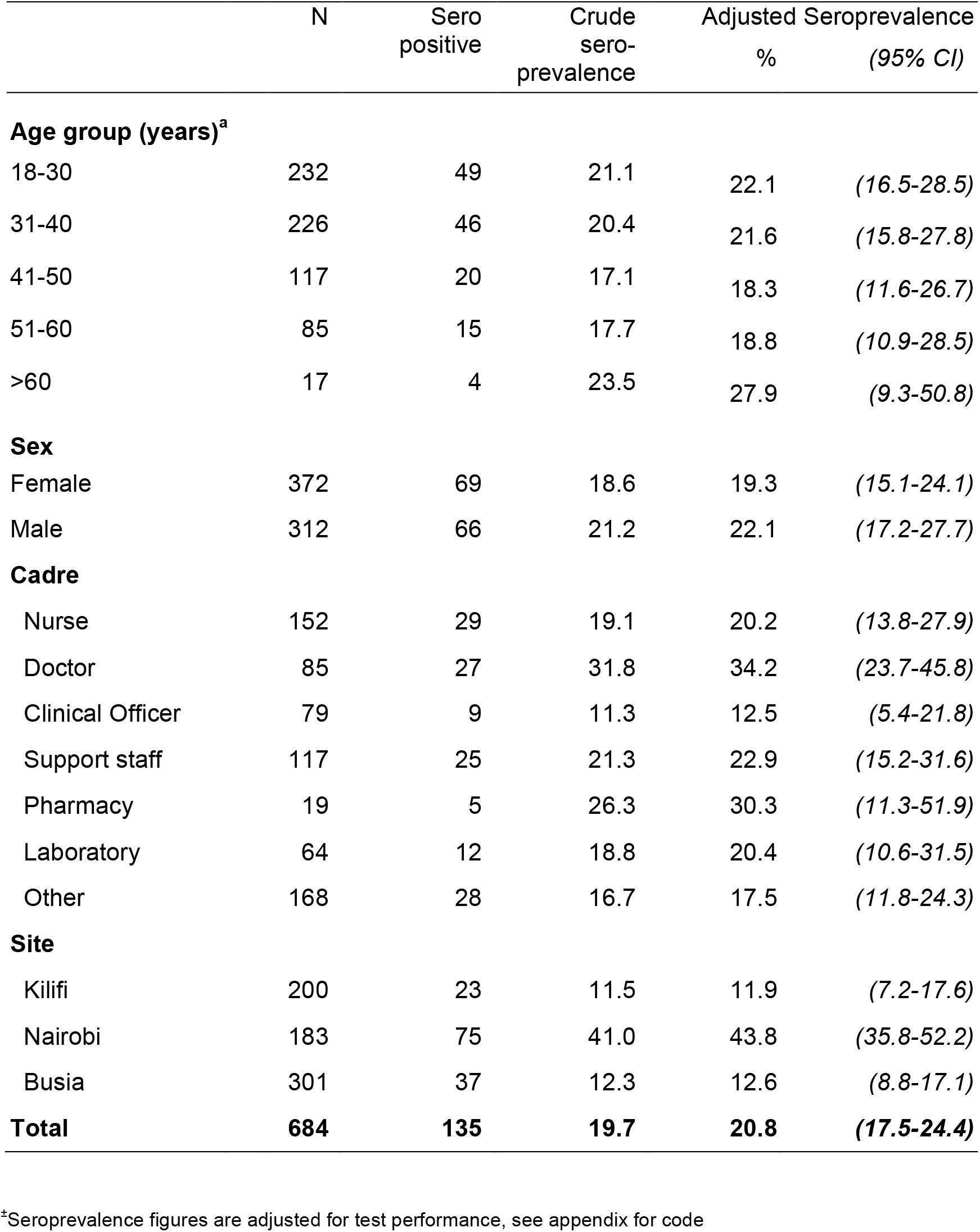
Seroprevalence of antibodies to SARS-CoV-2 by participant characteristics.

|  | N | Sero<br>positive | Crude<br>sero-<br>prevalence | Adjusted<br>% Seroprevalence | (95% CI) |
| --- | --- | --- | --- | --- | --- |
| **Age group (years)<sup>a</sup>** |  |  |  |  |  |
| 18-30 | 232 | 49 | 21.1 | 22.1 | (16.5-28.5) |
| 31-40 | 226 | 46 | 20.4 | 21.6 | (15.8-27.8) |
| 41-50 | 117 | 20 | 17.1 | 18.3 | (11.6-26.7) |
| 51-60 | 85 | 15 | 17.7 | 18.8 | (10.9-28.5) |
| >60 | 17 | 4 | 23.5 | 27.9 | (9.3-50.8) |
| **Sex** |  |  |  |  |  |
| Female | 372 | 69 | 18.6 | 19.3 | (15.1-24.1) |
| Male | 312 | 66 | 21.2 | 22.1 | (17.2-27.7) |
| **Cadre** |  |  |  |  |  |
| Nurse | 152 | 29 | 19.1 | 20.2 | (13.8-27.9) |
| Doctor | 85 | 27 | 31.8 | 34.2 | (23.7-45.8) |
| Clinical Officer | 79 | 9 | 11.3 | 12.5 | (5.4-21.8) |
| Support staff | 117 | 25 | 21.3 | 22.9 | (15.2-31.6) |
| Pharmacy | 19 | 5 | 26.3 | 30.3 | (11.3-51.9) |
| Laboratory | 64 | 12 | 18.8 | 20.4 | (10.6-31.5) |
| Other | 168 | 28 | 16.7 | 17.5 | (11.8-24.3) |
| **Site** |  |  |  |  |  |
| Kilifi | 200 | 23 | 11.5 | 11.9 | (7.2-17.6) |
| Nairobi | 183 | 75 | 41.0 | 43.8 | (35.8-52.2) |
| Busia | 301 | 37 | 12.3 | 12.6 | (8.8-17.1) |
| **Total** | **684** | **135** | **19.7** | **20.8** | **(17.5-24.4)** |

<sup>a</sup>Seroprevalence figures are adjusted for test performance, see appendix for code

Table 3 displays the results of univariable and multivariable logistic regression modeling testing associations between participant characteristics and seroprevalence. The only exposure variable that displayed a statistically significant association with seroprevalence in the multivariable model was site; HCWs in Kilifi (OR 0.2, 95% CI 0.1-0.3) and Busia (OR 0.2, 95% CI 0.1-0.4) were less likely to be seropositive compared to those in Nairobi.

**Table 3:** Univariable and multivariable analysis of factors associated with presence of antibodies to SARS-CoV-2.

| Characteristic | Univariable |  | Multivariable<sup>§</sup> |  |
| --- | --- | --- | --- | --- |
|  | OR | (95 % CI) | OR | (95% CI) |
| Sex |  |  |  |  |
| Female | 1.0 | – | 1.0 | – |
| Male | 1.18 | (0.81-1.71) | 1.13 | (0.75-1.72) |
| Age (per decade) | 1.00 | (0.98-1.01) | 0.99 | (0.98-1.01) |
| Site |  |  |  |  |
| Nairobi | 1.0 | – | – | – |
| Kilifi | 0.19 | (0.11-0.32) | 0.18 | (0.10-0.33) |
| Busia | 0.20 | (0.13-0.32) | 0.21 | (0.13-0.36) |
| Working in Covid unit | 0.33 | (0.12-0.94) | 0.51 | (0.17-1.55) |
| Symptoms at sample collection | 1.98 | (1.08-3.63) | 1.34 | (0.78-2.30) |
| Chronic illness | 2.49 | (0.88-6.97) | 0.91 | (0.30-2.72) |
| Cadre |  |  |  |  |
| Nurse | 1.0 | – | 1.0 | – |
| Doctor | 1.97 | (1.07-3.63) | 1.20 | (0.61-2.35) |
| Clinical Officer | 0.55 | (0.24-1.21) | 0.97 | (0.41-2.30) |
| Support staff | 1.15 | (0.63-2.09) | 1.56 | (0.80-3.07) |
| Pharmacy | 1.51 | (0.51-4.54) | 1.50 | (0.45-4.97) |
| Laboratory | 0.98 | (0.46-2.06) | 1.45 | (0.64-3.27) |
| Other | 0.85 | (0.48-1.50) | 0.97 | (0.53-1.81) |

<sup>§</sup>Adjusted for all variables in table

Professional cadre, age and sex were not associated with seroprevalence in both univariable and multivariable analyses. Site-specific analyses also did not reveal any association between seroprevalence and professional cadre (Table S1).

## DISCUSSION

We report results of a SARS-CoV-2 seroprevalence study conducted among HCWs in 3 counties in Kenya. We found an overall seroprevalence of SARS-CoV-2 antibodies of 20.8% (95% CI 17.5-24.4%). There were significant differences in seroprevalence associated with hospital region, but no differences associated with professional cadre.

Our estimates of seroprevalence are higher than what was found in most of studies from Africa that have been published to date, all of which were conducted in urban areas^6-8, 16^ and had a pooled seroprevalence of 8.2% (95% CI 0.8-22.3)^17^. We conducted our study during and shortly after the first wave of the epidemic in Kenya^4^, while the previous studies in Africa were conducted relatively early in the epidemic. Our estimates are similar to those observed among HCWs in several high-income countries at the peak of their first wave of the epidemic^17^.

Consistent with other studies conducted in Kenya^4, 14, 18-20^, we found significant differences in seroprevalence by region. HCWs in urban Nairobi had significantly higher seroprevalence than those in Busia and Kilifi, which are rural counties. Studies in Spain and India have also shown significant regional differences, with higher seroprevalence in urban areas, such as Madrid and New Delhi, compared to rural areas^21, 22^. However, even in the rural counties in Kenya, HCWs had seroprevalence estimates that were similar to those in HCWs in urban areas in Spain^23^, USA^24^ and Malawi^6^.

We found no differences in seroprevalence by professional category even when the analyses were stratified by study site. The absence of differences in seroprevalence by cadre in the presence of significant differences by geographical region suggests that community transmission could be playing a bigger role than workplace exposure. In studies of HCWs conducted in the UK, the incidence of infection mirrored that seen in the community^2, 25^. This suggests that efforts to suppress community transmission are likely to reduce infections among HCWs.

The results of this study provide further evidence that there has been significant undocumented transmission of the SARS-CoV-2 virus within Kenya. Additional evidence of significant undocumented transmission in Kenya derives from (1) two studies of seroprevalence among blood transfusion donors^14, 18^; (2) a study of truck drivers and their assistants conducted at the same time as this survey in Kilifi and Busia that found a seroprevalence of 42%^19^, and; (3) In a study of antenatal clinic attendees, seroprevalence was 50% at Kenyatta National Hospital in August 2020, and 11% at Kilifi County Hospital in November 2020^20^.

A particular strength of this study is that we conducted it in several sites, which enabled us to detect a significant burden of infection among HCWs in rural parts of the country. Another strength is that we used an assay that was validated using both local and external samples and which performed well in a WHO-sponsored international standardization study^26^. Although we adjusted our figures using Bayesian modelling to take into account assay performance, the reported seroprevalence could still be underestimated due to antibody waning^27^. The longitudinal phase of the current study will help address this issue. Another possible reason for underestimation of the prevalence in our study would be spectrum bias^28^ since the samples that we used in validating the assay, although derived from the local population, these individuals were not necessarily the same as the HCWs that participated in the present survey.

Our study had several limitations. We did not perform genetic sequencing to establish the likely sources of infections among the HCWs, although as argued above, the data we obtained suggests that community transmission was the main driver of infections among the HCWs. The non-random selection of only a small proportion of the HCWs in Nairobi could have led to an overestimation of the seroprevalence if the HCWs sampled had an overrepresentation of individuals who had experienced symptoms in the past. However, this would have also resulted in a higher proportion of HCWs in Nairobi having positive results from previously conducted PCR tests, but we did not observe this. In addition a household survey found that 35% of the population in Nairobi had antibodies to SARS-CoV-2^29^, and the rural-urban difference in seroprevalence among HCWs that we observed was similar to what has been observed in other studies conducted in Kenya^14, 18-20^.

In conclusion, we found a high prevalence of antibodies to SARS-CoV-2 among HCWs in Kenya, with significant regional differences and no differences based on cadre. The results suggest that infection with SARS-CoV-2 among HCWs is driven more by background population levels of infection than workplace exposure and will be useful in informing measures to control the on-going pandemic.

## Data Availability

De-identified data are avaialble upon reasonable request

## Acknowledgments

We would like to thank all HCWs who participated in the study as well as the county health teams and the Kenya Paediatric Research Consortium (KEPRECON) that facilitated data collection for the study. We thank F. Krammer for providing the plasmids used to generate the RBD, spike protein, and CR3022 monoclonal antibody used in this work. Development of SARS-CoV-2 reagents was partially supported by the NIAID Centres of Excellence for Influenza Research and Surveillance (CEIRS) contract HHSN272201400008C. The COVID-19 convalescent plasma panel (NIBSC 20/118) and research reagent for SARS-CoV-2 Ab (NIBSC 20/130) were obtained from the NIBSC, UK. We also thank the WHO SOLIDARITY II network for sharing of protocols and for facilitating the development and distribution of control reagents. This manuscript was written with the permission of the Director, KEMRI-CGMRC.

## Funding

This project was funded by the Wellcome Trust (grants 220991/Z/20/Z and 203077/Z/16/Z), the Bill and Melinda Gates Foundation (INV-017547), and the Foreign Commonwealth and Development Office (FCDO) through the East Africa Research Fund (EARF/ITT/039) and is part of an integrated programme of SARS-CoV-2 serosurveillance in Kenya led by KEMRI Wellcome Trust Research Programme. ***For the purpose of Open Access, the author has applied a CC-BY public copyright licence to any author accepted manuscript version arising from this submission***. A.A. is funded by a DFID/MRC/NIHR/Wellcome Trust Joint Global Health Trials Award (MR/R006083/1), J.A.G.S. is funded by a Wellcome Trust Senior Research Fellowship (214320) and the NIHR Health Protection Research Unit in Immunisation, I.M.O.A. is funded by the United Kingdom’s Medical Research Council and Department For International Development through an African Research Leader Fellowship (MR/S005293/1) and by the NIHR-MPRU at UCL (grant 2268427 LSHTM). G.M.W. is supported by a fellowship from the Oak Foundation. C.N.A. is funded by the DELTAS Africa Initiative [DEL-15-003], and the Department for International Development and Wellcome (220985/Z/20/Z). S.U. is funded by DELTAS Africa Initiative [DEL-15-003], L.I.O-O. is funded by a Wellcome Trust Intermediate Fellowship (107568/Z/15/Z).

## Conflicts of Interest/Disclosures

None of the authors have any conflicts of interest or disclosures to report.

## Supplementary Appendix

### 1. Data collection form

#### SARS CoV-2 surveillance in HCW

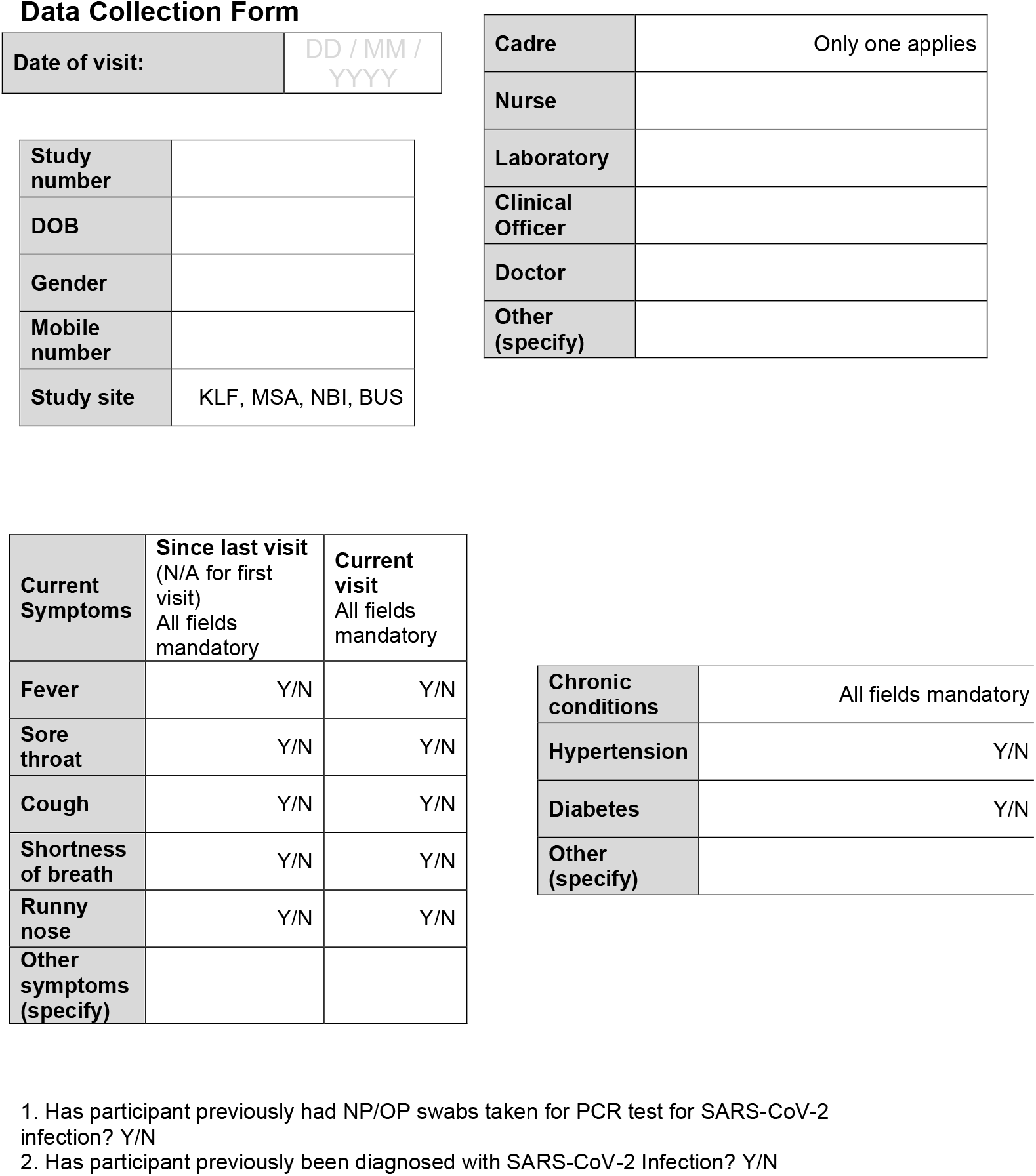

### 2. Stan code for Bayesian adjustment of prevalence estimates to account for test performance

~~~
data {
 int N;
 int N_se;
 int N_sp;
 int y;
 int x;
 int z;
}
parameters {
 real<lower=0,upper=1> p;
 real<lower=0,upper=1> se;
 real<lower=0,upper=1> sp;
}
transformed parameters {
 real<lower=0,upper=1> p_obs;
 p_obs = se * p + (1 - sp) * (1 - p);
}
model {
 //priors
 p ∼ beta(1, 1);
 se ∼ beta(1, 1);
 sp ∼ beta(1, 1);
 //likelihood
 y ∼ binomial(N, p_obs);
 x ∼ binomial(N_se, se);
 z ∼ binomial(N_sp, sp);
}
~~~

#### Data

Data:
|  | KNH | KCH | Busia | Nurse | Doctor | Clinical<br>Officer | Pharm<br>acy | Support<br>staff | Other |
| --- | --- | --- | --- | --- | --- | --- | --- | --- | --- |
| y | 75 | 23 | 37 | 29 | 27 | 9 | 5 | 25 | 28 |
| x | 166 | 166 | 166 | 166 | 166 | 166 | 166 | 166 | 166 |
| z | 901 | 901 | 901 | 901 | 901 | 901 | 901 | 901 | 901 |
| N | 183 | 200 | 301 | 152 | 85 | 79 | 19 | 117 | 162 |
| N_se | 179 | 179 | 179 | 179 | 179 | 179 | 179 | 179 | 179 |
| N_sp | 910 | 910 | 910 | 910 | 910 | 910 | 910 | 910 | 910 |

**Table S1:** Multivariable analysis of factors associated with presence of antibodies to SARS-CoV-2 in Nairobi, Kilifi and Busia.

| Characteristic | Nairobi |  | Kilifi |  | Busia |  |
| --- | --- | --- | --- | --- | --- | --- |
|  | OR | (95% CI) | OR | (95% CI) | OR | (95% CI) |
| Sex |  |  |  |  |  |  |
| Female | 1.0 | – | 1.0 | – | 1.0 | – |
| Male | 1.11 | (0.60-2.05) | 0.69 | (0.26-1.81) | 1.54 | (0.74-3.19) |
| Age category<sup>§</sup> | 0.96 | (0.73-1.26) | 0.82 | (0.52-1.30) | 0.98 | (0.69-1.39) |
| Work in Covid Unit<sup>±</sup> | – | – | – | – | 0.49 | (0.16-1.51) |
| Cadre |  |  |  |  |  |  |
| Nurse | 1.0 | – | 1.0 | – | 1.0 | – |
| Doctor | 0.87 | (0.38-1.98) | 1.40 | (0.17-6.41) | 4.62 | (0.84-25.3) |
| Clinical Officer | 0.44 | (0.04-4.60) | 1.18 | (0.29-4.89) | 1.68 | (0.34-8.37) |
| Support staff | 1.35 | (0.47-3.86) | 1.25 | (0.24-6.43) | 2.72 | (0.76-9.75) |
| Lab & Pharmacy | 1.24 | (0.37-4.14) | 2.44 | (0.46-12.96) | 1.89 | (0.50-7.11) |
| Other | 0.60 | (0.25-1.44) | 1.58 | (0.38-6.56) | 1.65 | (0.48-5.74) |

<sup>§</sup> Per decade increase in age
<sup>±</sup> Only applied to HCWs in Busia. None of the HCWs in Nairobi and Kilifi worked in a Covid isolation unit

**Figure S1:**
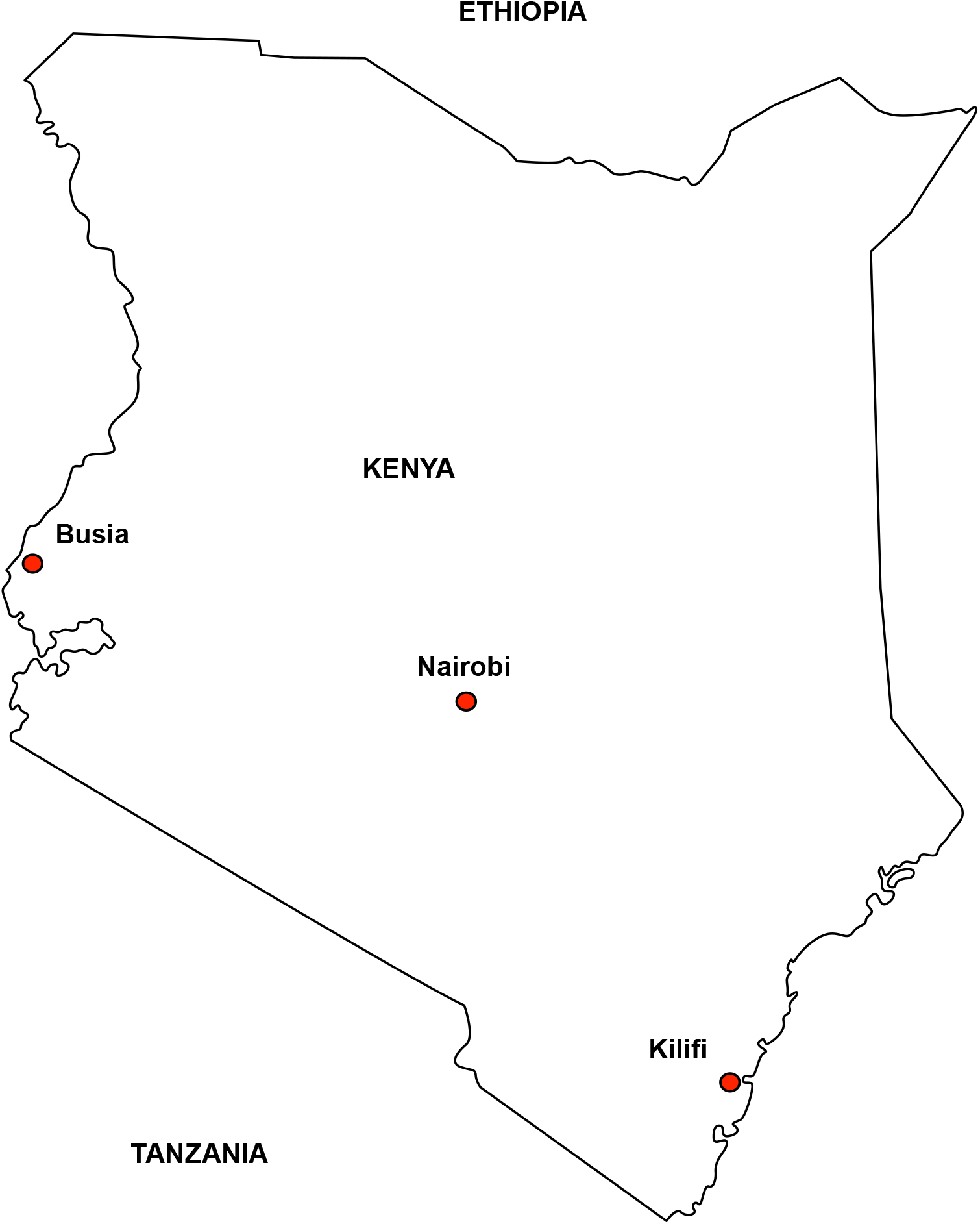
Study locations.

## Notes

### Competing Interest Statement

The authors have declared no competing interest.

### Author Declarations

Kenya Medical Research Institute Scientific and Ethics Review Unit (KEMRI/SERU/CGMR-C/203/4085)

